# Premature deaths attributable to the consumption of ultra-processed foods: a comparative assessment modelling study in eight countries

**DOI:** 10.1101/2023.10.05.23296603

**Authors:** Eduardo A. F. Nilson, Felipe Mendes Delpino, Carolina Batis, Priscila Pereira Machado, Jean-Claude Moubarac, Gustavo Cediel, Camila Corvalan, Gerson Ferrari, Fernanda Rauber, Euridice Martinez-Steele, Maria Laura da Costa Louzada, Renata Bertazzi Levy, Carlos A. Monteiro, Leandro F.M. Rezende

**Affiliations:** Center of Epidemiological Research in Nutrition and Health, University of Sao Paulo, Brazil; Oswaldo Cruz Foundation (Fiocruz/Brasilia), Brasília, Brazil; Postgraduate Program in Nursing, Federal University of Pelotas, Brazil; Health and Nutrition Research Center, National Institute of Public Health, Cuernavaca, Mexico; Deakin University. Institute for Physical Activity and Nutrition. Melbourne, Australia; Department of Nutrition, Faculty of Medicine, Université de Montréal, Canada; School of Nutrition and Dietetics, University of Antioquia, Medellin, Colombia; CIAPEC, Institute of Nutrition and Food Technology (INTA), University of Chile. Chile; Universidad de Santiago de Chile (USACH), Escuela de Ciencias de la Actividad Física, el Deporte y la Salud, Santiago, Chile; Department of Preventive Medicine, Faculdade de Medicina FMUSP, Universidade de São Paulo, Sao Paulo, Brazil; Department of Nutrition, School of Public Health, University of São Paulo, São Paulo, Brazil; Center for Epidemiological Studies in Health and Nutrition, University of São Paulo, São Paulo, Brazil; Department of Preventive Medicine, Escola Paulista de Medicina, Universidade Federal de São Paulo, Sao Paulo, Brazil

## Abstract

**Background:** Ultra-processed foods (UPFs) are becoming dominant in the global food and supply. Prospective cohort studies have found an association between UPF dietary pattern and increased risk of several non-communicable diseases and all-cause mortality. In this study, we (1) estimated the risk of all-cause mortality associated for each 10% increase in the share of UPF consumption in the total energy intake; (2) estimated the population attributable fractions (PAF) and the total number of premature deaths attributable to the consumption of UPF in adults (30-69 years) from 8 selected countries.

**Methods:** First, we performed a dose-response meta-analysis of observational cohort studies assessing the association between UPFs dietary pattern and all-cause mortality. As we found evidence of linearity, we estimated the pooled RR (and its 95% CI) for all-cause mortality per each 10% increment in the % UPF. Then, we estimated the population attributable fraction (PAF) of premature all-cause mortality attributable to UPF in 8 selected countries with relatively low (Colombia and Brazil), intermediate (Chile and Mexico), and high (Australia, Canada, UK, and US) UPF consumption.

**Results:** We found a linear dose-response association between UPF intake and all-cause mortality, with a 2.7% increased risk of all-cause mortality per 10% increase in the % UPF. Considering the magnitude of the association between UPFs intake and all-cause mortality, and the dietary share of UPF in each of the 8 selected countries, we estimated that 4% (Colombia) to 14% (United Kingdom and United States) of premature deaths were attributable to UPF intake.

**Conclusions:** Our findings support that UPF intake contributes significantly to the overall burden of disease in many countries and its reduction should be included in national dietary guideline recommendations and addressed in public policies.

## INTRODUCTION

Ultra-processed foods (UPFs), as defined by the Nova classification^1^, are becoming dominant in the global food supply^2^. UPFs account for over half of the energy content of diets in some high-income countries, such as US and UK^3,4,5^. Although UPFs remains lower in low-and middle-income countries, there is evidence that its consumption has increased rapidly worldwide over the last decades^6^. Prospective cohort studies have found an association between UPF dietary pattern and increased risk of several non-communicable diseases and all-cause mortality^7,8,9^. To our knowledge, however, the shape of the dose-response relationship between energy share of UPFs and all-cause mortality remains unknown. In addition, the burden of deaths attributable to UPF consumption have not yet been modelled in countries with different dietary share of UPF. In this study, we (1) estimated the risk of all-cause mortality associated for each 10% increase in the share of UPF consumption in the total energy intake;; (2) estimated the population attributable fractions (PAF) and the total number of premature deaths (herein defined as deaths between 30 to 69 years of age) attributable to the consumption of UPF in adults (30-69 years) from 8 selected countries with relatively low (Colombia and Brazil), intermediate (Chile and Mexico), and high (Australia, Canada, UK, and US) UPF consumption.

## METHODS

### Meta-analysis for the association between ultra-processed foods consumption and all-cause mortality

First, we performed a dose-response meta-analysis of observational cohort studies assessing the association between UPFs dietary pattern and all-cause mortality. Studies were selected based on recently published systematic reviews on high vs. low UPF dietary pattern and all-cause mortality ^7,10,11^. We included only studies that assessed UPF intake according to Nova classification and excluded studies assessing UPF as individual food items (e.g. sugar-sweetened beverages). In the previously published meta-analyses^7,10,11^, 10 prospective studies examined the association between UPF dietary pattern and all-cause mortality ^12,13,14,15,16,17,18,19,20,21^. whether they could provide estimates after converting UPF to % of total energy intake. We obtained information from 7 studies^12,13,14,15,16,17,18^. We extracted the maximally-adjusted relative risks (RR) and 95% confidence intervals (CI) for all-cause mortality for each category of % UPF to total energy intake (e.g., quartiles of % UPF) and considered the mean or medium value of % of UPF in a given category (aka dose). For the highest category of UPF intake (e.g., 4th quartile), we considered the dose as the lowest value of % UPF in the category. We also extracted the number of participants and deaths in each category of % UPF. We ran a random-effects dose-response model using generalized least squares for trend estimation of summarized dose-response data. As we found evidence of linearity, we estimated the pooled RR (and its 95% CI) for all-cause mortality per each 10% increment in the % UPF. The robustness of the results was established by eliminating each study one by one from the meta-analysis and recalculating the summary estimate (the ‘leave one out’ approach).

### Premature deaths attributable to the consumption of ultra-processed foods in selected countries

We estimated the population attributable fraction (PAF) of premature (30-69 years) all-cause mortality attributable to UPF in 8 selected countries with relatively low (Colombia and Brazil), intermediate (Chile and Mexico), and high (Australia, Canada, UK, and US) UPF consumption, using the following equation:

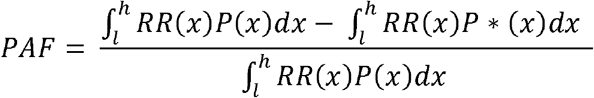

Where x denotes the level exposure to %UPF (0 to 100%); RR is the RR function (log-linear) for all-cause mortality as reported in the previous section; P(x) is the % of UPF intake in each country and P*(x) is the theoretical minimum risk exposure level (assumed as 0% of UPF intake); dx denotes that the integration was done with respect to x, and l and h are the integration boundaries.

The mean and 95% CI of % UPFs intake in each country was estimated from the most recent nationally-representative dietary surveys using 24-hour recall data: Brazil (2017– 18), Chile (2010), Colombia (2015), Mexico (2016), Australia (2011–12), the United Kingdom (2016-19), Canada (2015), and the United States of America (2017-18). Information on the dietary surveys has been reported elsewhere ^22^.

For each country, we retrieved the total number of premature deaths that occurred in the same years of the national dietary survey from the Global Burden of Disease Study. Monte Carlo simulation with 5,000 iterations was used to estimate the uncertainty of the PAF attributable to the consumption of UPF.

Data analyses were performed in Stata Version 17, Microsoft Excel and Ersatz.

## RESULTS

The main results extracted from each of the 7 prospective cohort studies included in the meta-analysis are displayed in Table 1. The dose-response meta-analysis for the association between the UPF consumption (% energy intake) and all-cause mortality including 239,982 participants and 14,779 deaths is presented in Figure 1. The pooled RR for each 10% increase in % UPF energy intake was 1.027 (95 % CI 1.017 to 1.037; P < 0.0001). Leave-one-out sensitivity analysis provided results consistent with the main analysis (Table S1).

**Table 1:**
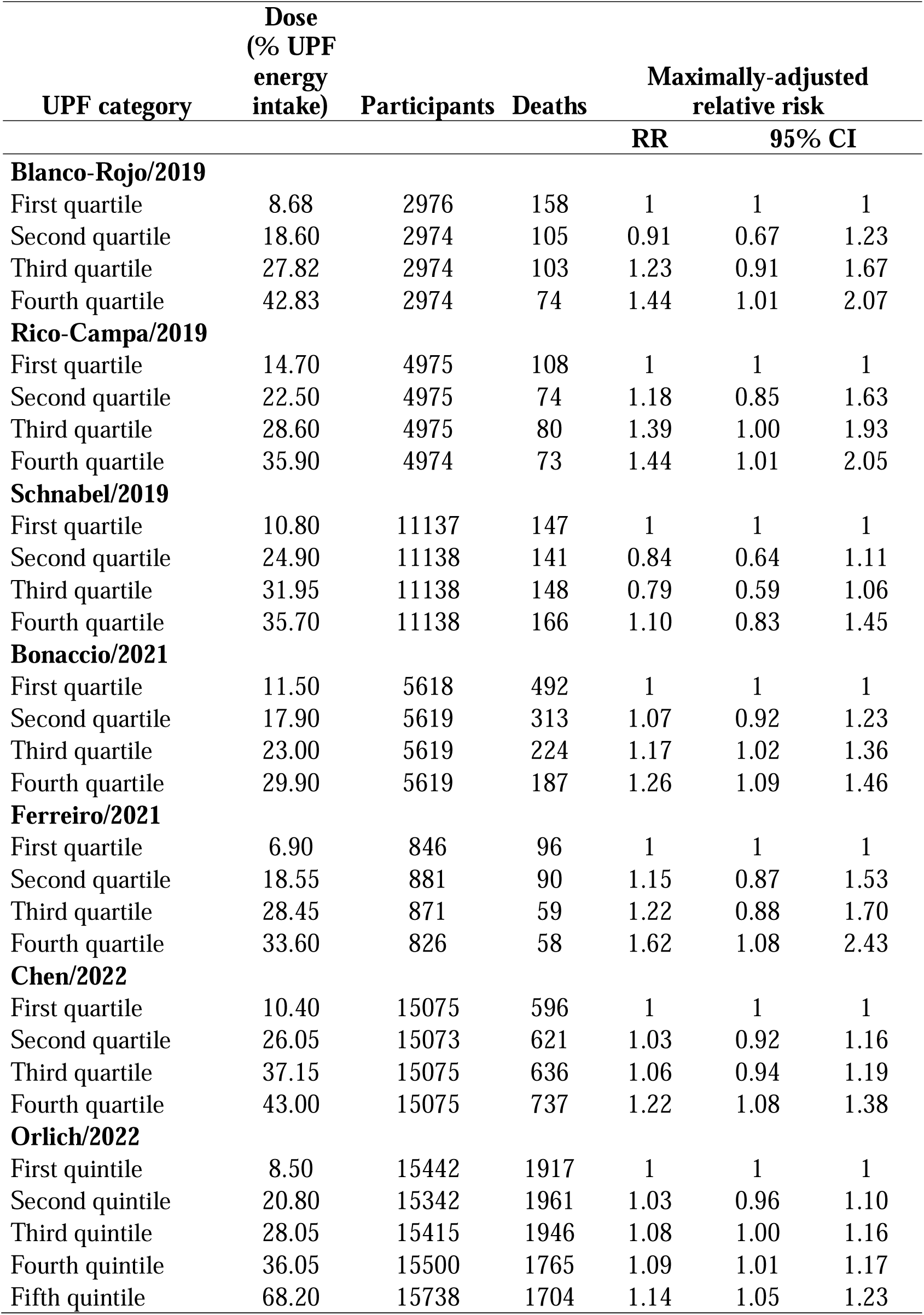
Main results extracted from 7 prospective cohort studies included in the dose-response meta-analysis.

**Figure 1:**
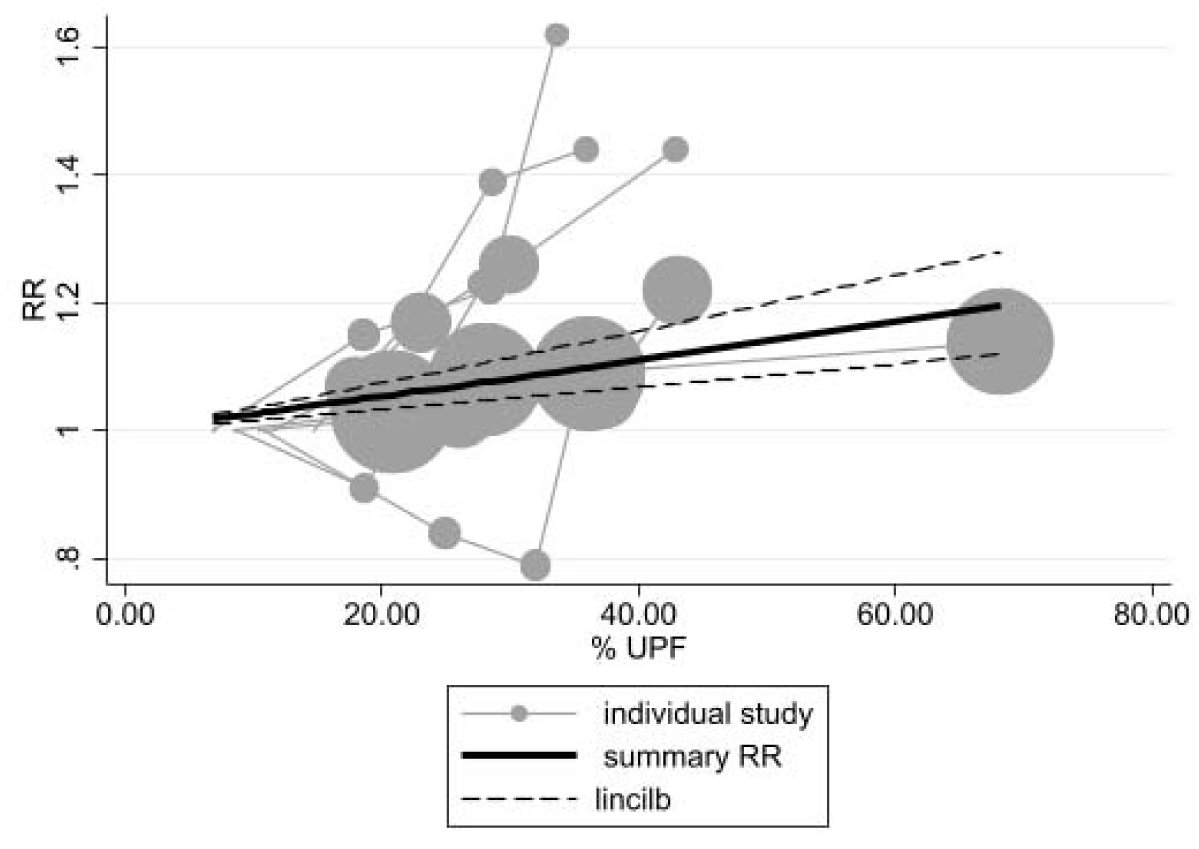
Dose-response meta-analysis for the association between the UPF consumption (% energy intake) and all-cause mortality from 7 prospective cohort studies including 239,982 participants and 14,779 deaths

In the 8 selected countries, PAFs ranged widely according to the consumption of UPF, ranging from 3.9% in Colombia, where UPF represents 15% of total energy intake, to almost 14% in the UK and the US, where UPF represents around 54% of total energy intake. The absolute number of premature deaths per year attributable to UPF (i.e., which is influenced by PAF, number of premature deaths and population size), ranged from nearly 2 thousand in Chile to over 124 thousand in the US (Table 2).

**Table 2:** Premature deaths attributable to the consumption of ultra-processed foods in 8 countries.

| Country, year | UPF intake<br>(% of total energy) |  |  | Population<br>Attributable Fraction |  |  | Premature Deaths |  |  |  |
| --- | --- | --- | --- | --- | --- | --- | --- | --- | --- | --- |
|  |  |  |  |  |  |  | Total | Attributable to UPF intake |  |  |
|  | UPF | 95% CI |  | % | 95% CI |  | Number | Number | 95% CI |  |
| **Colombia, 2015** | 15.0 | 14.2 | 15.7 | 3.9 | 2.5 | 5.4 | 72,940 | 2,813 | 1,860 | 3,906 |
| **Brazil, 2017-18** | 17.4 | 17.1 | 17.7 | 4.5 | 3.0 | 6.2 | 556,696 | 25,296 | 16,887 | 34,750 |
| **Chile, 2010** | 22.8 | 21.6 | 24.0 | 5.7 | 3.8 | 8.0 | 32,607 | 1,874 | 1,231 | 2,610 |
| **Mexico, 2016** | 24.9 | 22.6 | 27.1 | 6.3 | 4.1 | 8.8 | 272,293 | 17,110 | 11,184 | 23,852 |
| **Australia, 2011-13** | 37.5 | 36.8 | 38.2 | 9.4 | 6.3 | 12.8 | 34,994 | 3,277 | 2,204 | 4,472 |
| **Canada, 2015** | 43.7 | 42.7 | 44.7 | 10.9 | 7.3 | 14.8 | 70,994 | 7,735 | 5,175 | 10,527 |
| **UK, 2016-19** | 52.7 | 51.6 | 63.8 | 13.9 | 9.1 | 18.7 | 128,743 | 17,894 | 11,729 | 24,132 |
| **US, 2017-18** | 54.5 | 52.8 | 56.1 | 13.7 | 9.1 | 18.8 | 906,795 | 124,107 | 82,804 | 170,801 |

## DISCUSSION

In this study, we found a linear dose-response association between UPF intake and all-cause mortality, with a 2.7% increased risk of all-cause mortality per 10% increase in the % UPF. Considering the magnitude of the association between UPFs intake and all-cause mortality, and the dietary share of UPF in each of the 8 selected countries, we estimated that 4% (Colombia) to 14% (United Kingdom and United States) of premature deaths were attributable to UPF intake. Our findings support that UPF intake contributes significantly to the overall burden of disease in many countries and its reduction should be included in national dietary guideline’s recommendations and addressed in public policies. Limitations of our meta-analysis include the limited number of cohort studies that evaluated the association between UPF intake and all-cause mortality. Limitations of the model include the possibility of residual confounding of the RR estimate and the failure to capture potential time lag between dietary changes and mortality. These findings highlight that UPF consumption represents a relevant public health concern and should be addressed by evidence-based recommendations and policies.

## Competing interests

None to declare.

### Acknowledgements

We would like to thank all the researchers who, upon request, sent information on the original studies included in our dose-response meta-analysis

### Data Availability

All data produced in the present work are contained in the manuscript

## SUPPLEMENTARY MATERIAL

**Table S1:**
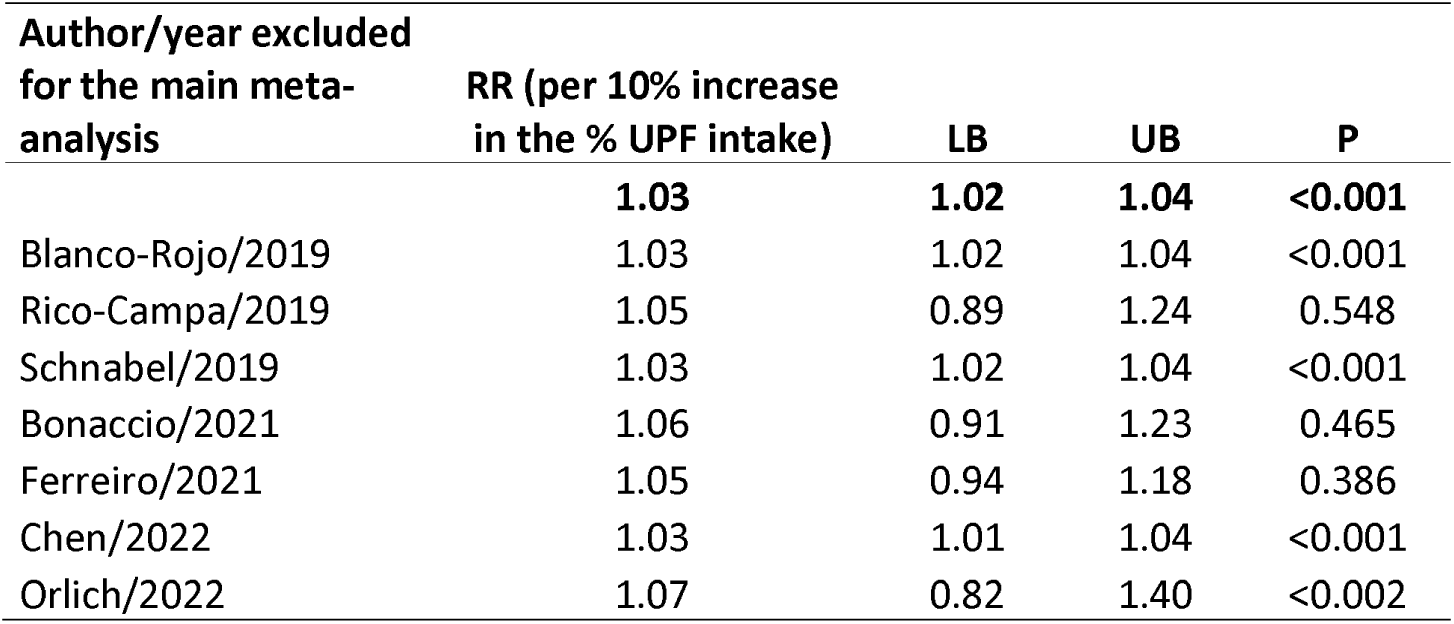
Sensitivity analysis using leave-one-out approach for the association between the consumption of ultra-processed foods and all-cause mortality.

